# Study the Prevalence of Multidrug resistant bacteria from air samples collected from Mumbai & Suburban areas

**DOI:** 10.1101/2024.08.22.24312400

**Authors:** Janvi Shah, Sourav Ghosh

## Abstract

**Introduction:** Air comprises mixture of airborne bacteria & air pollutants in near surface of atmosphere. Air pollution is one of the major threat around the world due to change in physical and chemical parameters because of natural and anthropogenic activities; leading to climate change.

**Materials and Methods:** A systematic analysis of AQI included air pollutants PM_2.5_, PM_10_, CO, SO_2_, NO_2_, O_3_, NH_3_ using SAMEER app developed by CPCB and meteorological parameters including wind speed and relative humanity. The overuse of antibiotics since last two decades has lead pathogenic bacteria gaining resistance.

**Results:** The study was carried out to identify the prevalence of MDR bacteria in air from 8 different spatial variations of Mumbai & Suburban areas. 32 isolates from air were collected by settle plate technique. Enumeration and characterization of bacteria was performed. Antibiotic susceptibility test against 10 antibiotics; test showed that all bacterial isolates were sensitive to Tigecycline. The bacteria were highly resistant to Methicillin by 81.25 %. ARI of two isolates was 0.9 & MRI of Vasai, 0.625 was the highest. The prominent pollutant among 8 locations was found to be PM_2.5_, CO & PM_10_. PCA & hierarchical cluster analysis showed that MRI is not correlated to 8 parameters but, distinctly related to NH_3_.

**Conclusions:** The MDR bacteria travel from soil, plants, water, humans, places, inanimate objects into the air. Hence, it is necessary to maintain the sanitation, ecological balance & usage of antibiotics when necessary.

## Introduction

Millions of bacteria are found in our environment. Some bacteria are beneficial while some cause infections to humans and animals. Antibiotic drugs were discovered to treat these infectious bacteria in late 19^th^ century.^1^ Over the years, consumption of antibiotics has increased to treat diseases of humans and animals or as feed additive to stimulate growth of animals.^2^ Humans started to purchase antibiotics over-the-counter (OTC) without prescription. Bacteria got exposed to different dosage of drugs and became resistant. Over the generations, the bacteria kept on gaining resistance against different antibiotics. Antibiotic resistant bacteria have been observed with increasing frequency over the past several decades.^3^ To overcome the problem pharmaceutical companies kept on manufacturing new generation antibiotics.

The resistant bacteria are found all around in water, air, soil, food, life-forms and on inanimate objects. Hence, the humans and animals are more prone to get infected. The diversity of microbes in the air is on par with the diversity of microbes in the soil, a fertile environment for such life-forms. In fact, there is a large crossover between the microbes in the air of a city and the microbes in its soil.^4^ Antibiotic resistant bacteria gain resistance coming in contact with one organism to other by mode of mobile genetic elements like, plasmids, transposons and phages turns multi-drug resistant (MDR) or superbugs. Antimicrobial resistance in bacterial pathogens is a worldwide challenge associated with high morbidity and mortality.^5^ Air contains high concentrations of microbes and inhaled by humans, around 14 m^3^ air per day.^6^ Air contains allergens, particulate matter and different gases.^7^ These MDR bacteria can be attached to ambient particles and/or incorporated into water droplets of clouds, fog, and precipitation (*i.e.*, rain, snow, hail) in the air. Further, they are deposited back to earth’s surfaces via dry and wet deposition processes. They can possibly induce an effect on the diversity and function of aquatic and terrestrial ecosystems or impose impacts to human health through microbial pathogens dispersion. Their impact on ecosystem and public health are strong indications that air microbes can metabolically activate and adapted to the harsh atmospheric conditions.

Air pollution is one of the major concerns around the world emerged due to industrialization and emission of harmful gases like, methane, CFCs, etc. due to anthropogenic activities. This leads to climate change, ecological imbalance, environmental degradation, etc. They can also affect the atmospheric chemistry and physics, with important implications on meteorology and global climate.^8^

Some microbes present in the air can lead to bacterial infections/diseases. The prevalence of these microbes causing infections are of two types Gram positive (coagulase negative *Staphylococcus aureus, Staphylococcus aureus, Streptococci spp. & Enterococcus spp*.) and Gram negative bacteria (*Escherichia coli, Pseudomonas aeruginosa, Enterobacter aerogenes & Acinetobacter spp*.).^9^ Outdoor bacterial communities and their concentrations are also affected by geographical factors such as types of land use and their spatial distribution. The bacteria found in built environments therefore originate from any of the natural and man-made surroundings around humans. Therefore, to understand better the factors influencing bacterial concentrations and communities in built environments, we should study all the environments that humans contact as a single ecosystem.^10^ Air pollution causes severe impacts on humans leading to respiratory illness; asthma (children & adults), lung function, airway obstruction, obstructive pulmonary diseases in case of children and elderly.

This study was carried out across Mumbai and suburban areas to understand the prevalence of the multi-drug resistance bacteria present the air and its connection with air pollutants.

In the present scenario of pandemic caused by coronavirus, there is a linkage between acceleration of COVID19 and air pollution.^11^ During COVID19 lockdown in India, there was seen a decrease in air pollutants especially in particulate matter (PM) and improvement in the air quality.^12^ It’s important to do a microbial census to see what’s in the air we breathe. We may see very different populations of microbes in the air and that may have some health implications. Also, implementing plans for strict execution in controlling the air quality after lockdown.

The challenge of containing and controlling the threat of antibiotic resistant pathogens is daunting. A multi-disciplinary approach is required which must be applied, sustained and continuously refined. The key components for maintaining effective antimicrobial chemotherapy creating microbiome friendly drugs, continuous investment in new and innovative technologies; including diagnostics and vaccines and studying epigenetics to develop personalized medicines.

## Material and Methods

### Study area

The locations were all over Mumbai & Navi Mumbai; sampling was carried during winter season (December & January) (Table 1).

### Physiochemical parameters

The parameters were taken on the same day of the sampling like, wind speed (WS) & relative humidity (RH) from Google and AQI (Air quality index) was procured from SAMEER app by CPCB (Central Pollution Control Board) which included hourly estimation of PM_2.5_, PM_10_, NO_2_, SO_2_, CO, O_3_ & NH_3_.

### Collection of samples

The sampling was carried out during morning peak hours. The airborne bacterial samples were collected by settle plate technique/ surface impingement method^6^ on sterile Nutrient agar (all-purpose media), Salt Mannitol Agar & Endo Agar (Hi media). The plates were exposed to air for 30mins. Then the plates were incubated at Room temperature (RT/37°C) for 24 hours.

### Enumeration of bacteria

The bacterial colonies were enumerated after 24 hours of incubation at RT from Nutrient agar plate and colony forming unit per cubic meter (CFU/m^3^) of air was calculated using Omeliansky formula^13^:

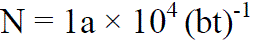

Where,

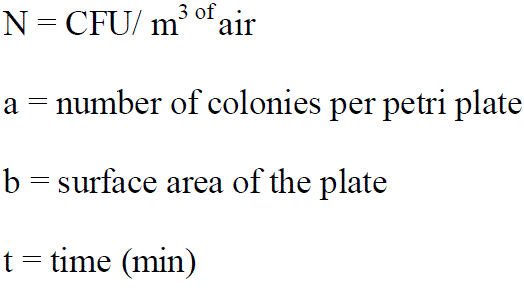

### Isolation of bacteria

The bacteria were isolated on selective media like, Salt Mannitol agar^14^ (Hi media M118) for Gram positive & Endo agar^15^ (Hi media M029) for Gram negative organisms.

### Identification of bacteria

Morphological characteristics of an isolated colony from Nutrient agar plate characters like size, colour, shape, margin, opacity, elevation, consistency and Gram staining were observed and recorded.^16^ Biochemical tests were performed according to the Gram character and referring Bergey’s Manual.^17^

### Antibiotic Susceptibility test (AST)

The antibiotic sensitivity for each of the isolate was evaluated by the method of Mishra et al.^18^ with few modifications. 1 ml of sterile Nutrient Broth (Hi media) was added into a 24 well coated microtitre plate (Tarson, 980030, Korea) to which 20 µl of 20 hour old test culture was added with 10 different antibiotic discs were placed into the wells to test and incubated at 37°C, for 24 hours and turbidity is checked. A negative and positive control was also maintained for comparison.

Ten different types antibiotic discs (Hi media) were chosen according to CLSI (Clinical & Laboratory Standards Institute) chart i.e. Ampicillin-25mcg (SD077-1VL), Ceftriaxone-30mcg (SD065-1VL), Chloramphenicol-30mcg (SD006-1VL), Cloxacillin-10mcg (SD143-1VL), Erythromycin-15mcg (SD083-1VL), Methicillin-10mcg (SD136-1VL), Norfloxacin-10mcg (SD057-1VL), Tigecycline-15mcg (SD278-1VL), Trimethoprim-5mcg (SD039-1VL) and Vancomycin-30mcg (SD045-1VL).

Multiple antibiotic Resistance Index (MRI) of each bacteria was calculated using the formula MRI = y / x, where ‘y’ represents the Actual Number of Resistance recorded from the observation, And ‘x’ represents the Total number of Antibiotics used in AST. Likewise, the Antibiotic Resistance Index (ARI) of each location was calculated using the formula ARI = y / nx, where ‘y’ represents the number of resistance recorded, ‘n’ represents the Population size and ‘x’ represents the number of Antibiotics used.^18^

### Data mining

The data curation of 8 locations with 11 parameters (AQI, PM_2.5_, PM_10_, NO_2_, SO_2_, CO, O_3_ & NH _3_, wind speed, relative humidity & MRI) was carried out. The values were obtained and raw data was standardized by Z-score method. Z-score is calculated by subtracting the values from mean value and divided by the standard deviation (SD) value.^19^

Formula:

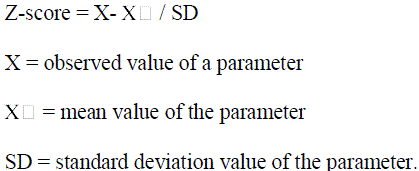

### Principal component analysis (PCA)

PCA develops a small set of uncorrelated components based on the scores on the variables. When one measures several variables, the correlation between each pair of variables can be arranged in a table of correlation coefficients between the variables. The existence of clusters of large correlation coefficients between subsets of variables suggests that the variables are related and could be measuring the same underlying dimension. These underlying dimensions are called components. A component is a linear combination of variables; it is an underlying dimension of a set of items.

The scree plot is one of the most common methods used for determining the number of components to extract. The scree plot is one of the procedures used in determining the number of factors to retain in factor analysis. In this procedure eigenvalues are plotted against their ordinal numbers and one examines where a break or a leveling of the slope of the plotted line occurs, the elbow. The components to the left of the point are considered significant. An eigenvalue is the amount of variance that a particular variable or component contributes to the total variance. This corresponds to the equivalent number of variables that the component represents. The concept of an eigenvalue is important in determining the number of components retained in principal component analysis. The eigenvalues chosen were ‘1’ and the components having a variance of less than ‘0.40’ were excluded.^20^

IBM SPSS Statistics V25 was used to perform PCA. KMO & Bartlett’s test of sphericity was used as correlation matrix. Rotation method used was Varimax. Scree plot was selected for identifying principal components.

### Cluster Analysis (CA)

Cluster analysis is a technique to form several cluster with similar values and bring about homogeneity (within cluster) and heterogeneity (between clusters). It is a basic tool for data analysis, involved detection, association, classification, clustering, summarization, etc. This method is accurate and fast compared to unsupervised techniques. Hierarchical Clustering is a type in which, the method creates a hierarchical decomposition of the given set of data objects. The tree of clusters formed is called as dendrograms. Their distances are measured between new cluster and each of old clusters.^21^

## Results and Discussion

The air is a mixture of gases, dust, allergens, pollutants, microbes, etc. The air samples were collected from 8 locations of Mumbai & Suburban areas in winter during peak timings (Figure 1), by settle plate method. AQI and 9 physiochemical parameters were recorded (Table 2). The airborne bacterial count (Table 3) of these areas was below average concentration i.e. ∼10^4^ CFU/m^3^.^8^ *Proteobacteria, Cyanobacteria, Firmicutes, Actinobacteria,* and *Bacteroidetes* were the dominant phyla among the total bacteria. *Cyanobacteria*, phototrophic bacteria, are active in atmosphere.^22^ Total number of 32 microbes was isolated and characterized; out of them 12 were Gram negative and 20 Gram positive.

**Figure 1.**
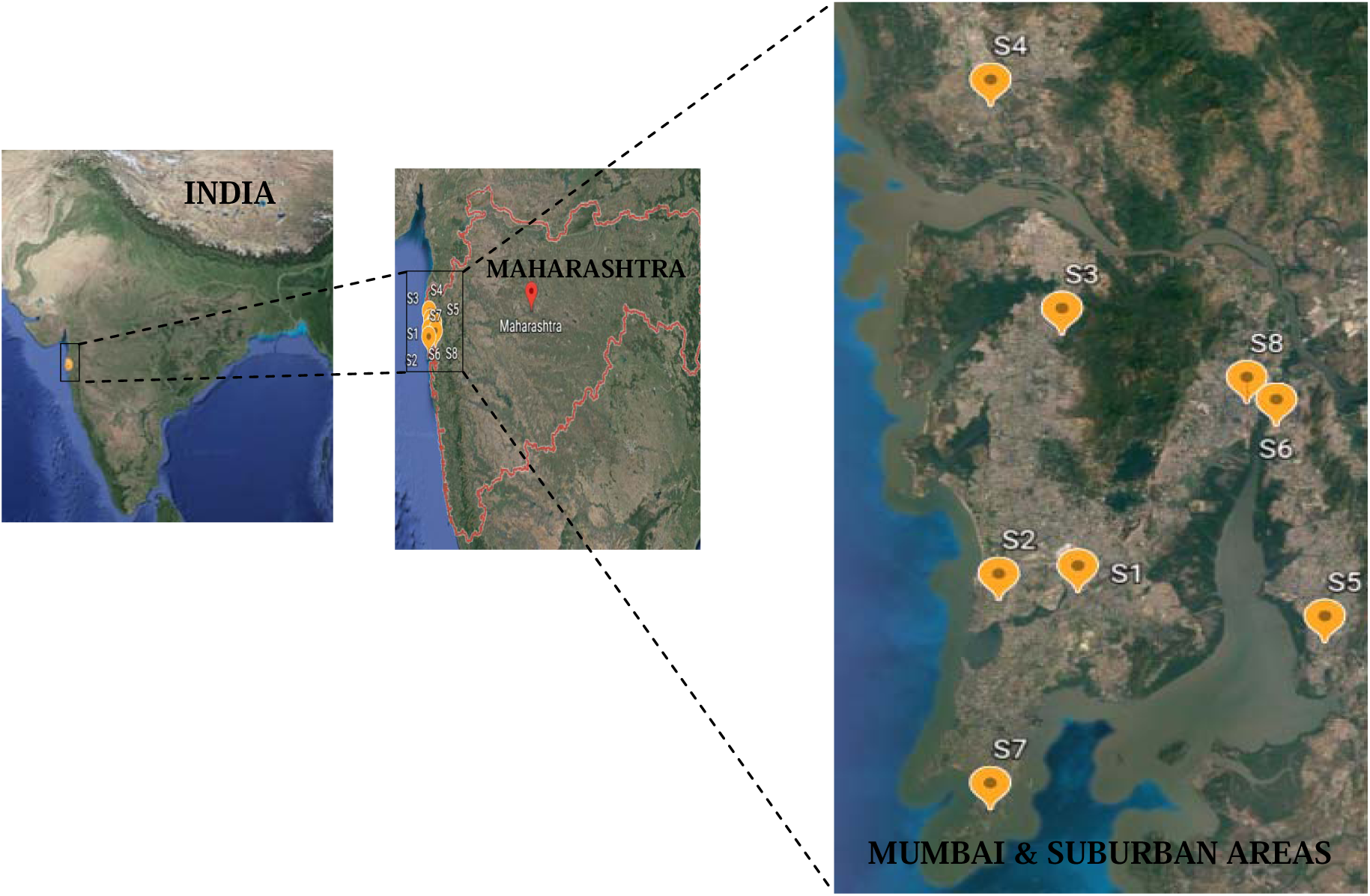
Google Earth map showing locations of sampling area.

AST of these isolates were tested against 10 antibiotics (Figure 2). All the isolates were sensitive to Tigecycline (100%) followed by Norfloxacin (87.5%), Chloramphenicol & Erythromycin (75%), Ampicillin (66%), Ceftriaxone & Vancomycin (62.5%), Cloxacillin (50%), Methicillin & Trimethoprim (18.75%). The isolates had highest resistant to Methicillin (81.25%) followed by Trimethoprim (68.75%), Cloxacillin (46.87%), Ceftriaxone (37.5%), Vancomycin (28.12%), Ampicillin (25%), Chloramphenicol (15.62%), Norfloxacin (9.37%) & Erythromycin (3.12%). Hence, instead of methicillin & trimethoprim any other broad spectrum antibiotic could be given for bacterial infection. The ARI of isolates S1_I2 and S4_I3 had highest index of 0.9. MRI of S4, 0.625 and S1, 0.475 isolates found to be the highest; followed by S8 and S3, 0.425 (Table 4). These locations consist of superbugs, those having higher MRI.

**Figure 2.**
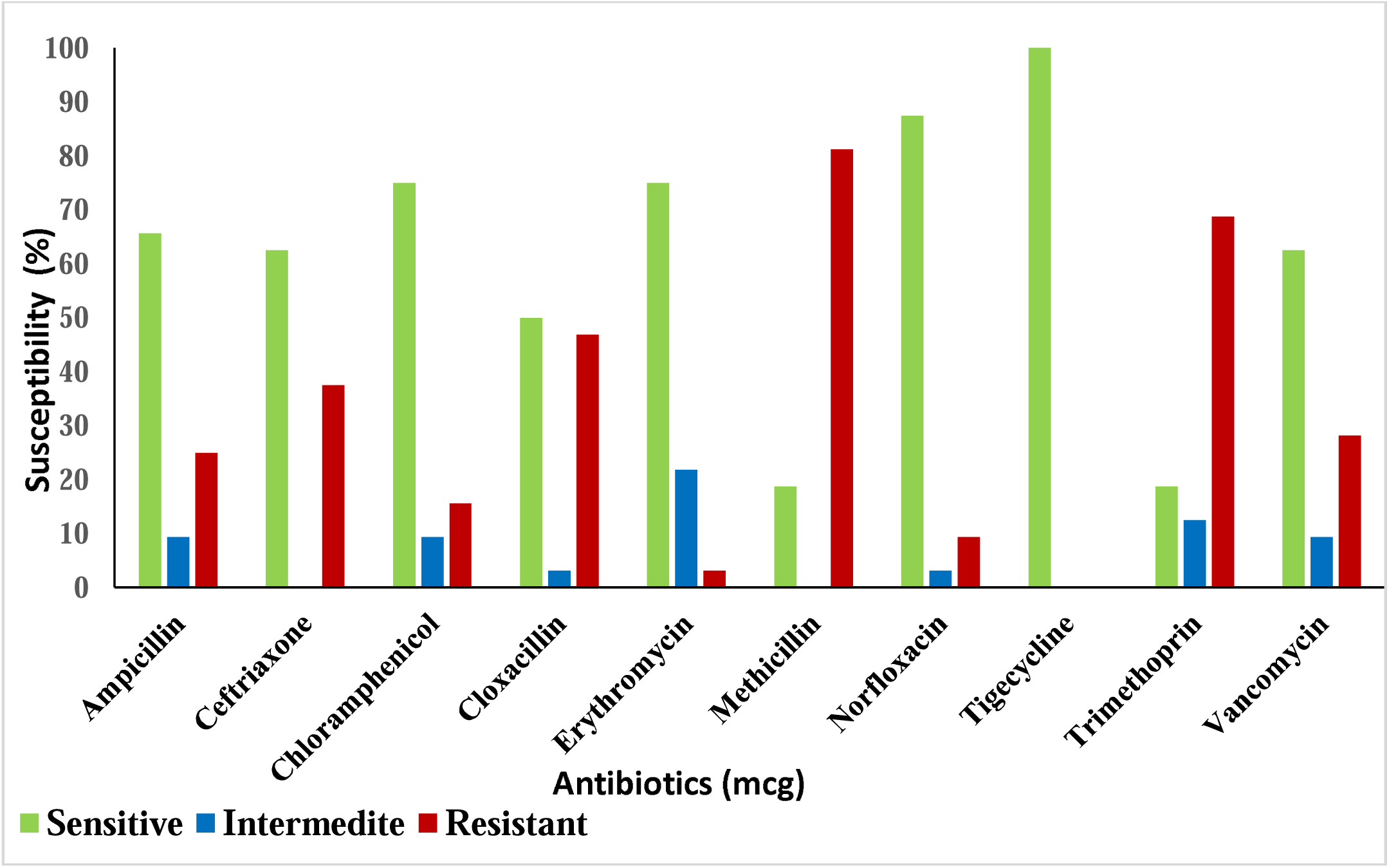
Antibiogram of percentage susceptibility of the isolates against 10 antibiotics.

The graphical representation of physiochemical parameters and MRI showed lower values of R-square (Figure 3). The 3 dimentional view of PCA showed that 9 parameters had interconnected relatedness and MRI found to be distinctly connected to NH_3_. Hence, the multidrug resistance of bacteria is independent of the physiochemical parameters (Figure 4).

**Figure 3.**
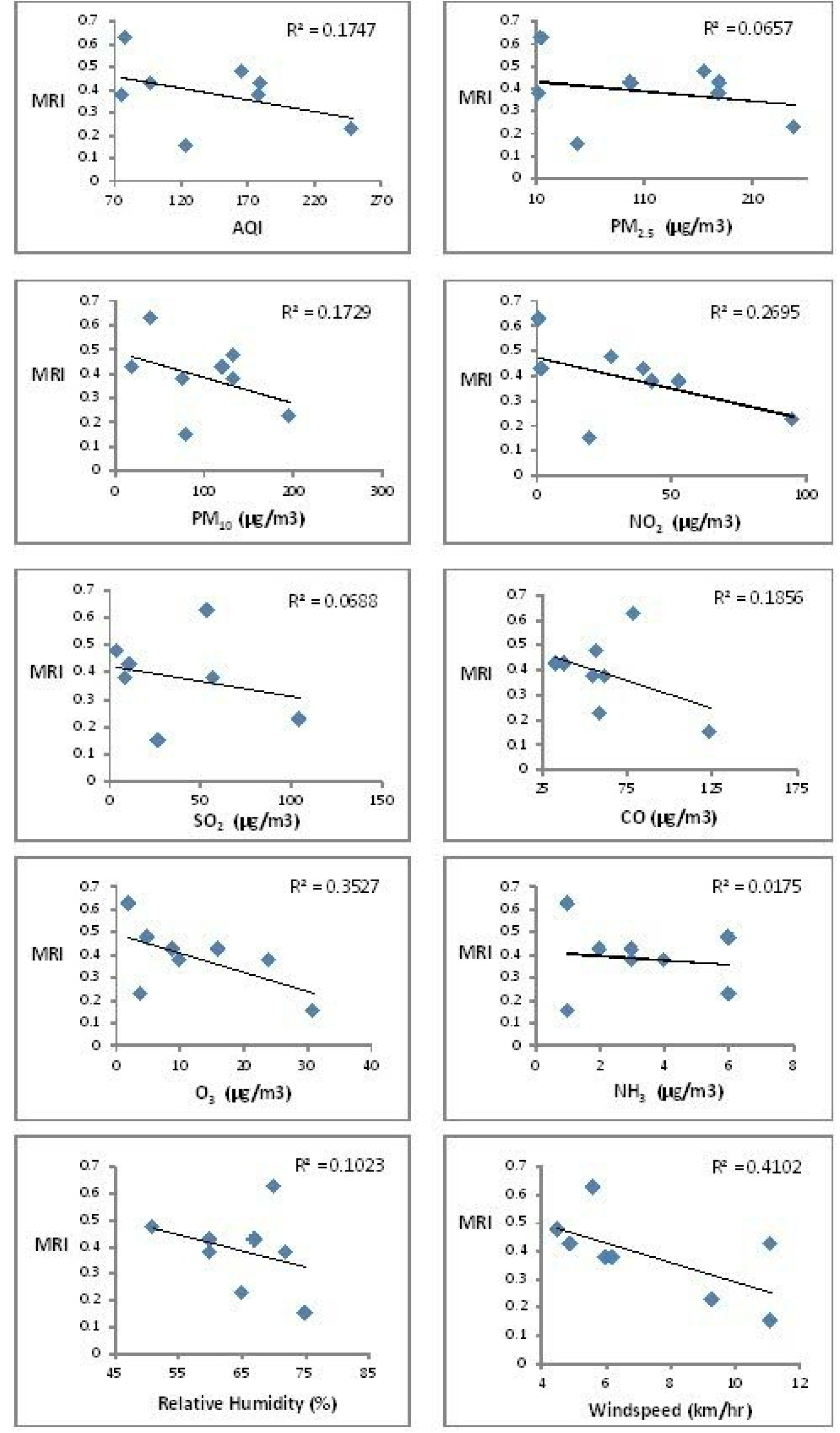
Graphical representation of physiochemical parameters v/s MRI.

**Figure 4.**
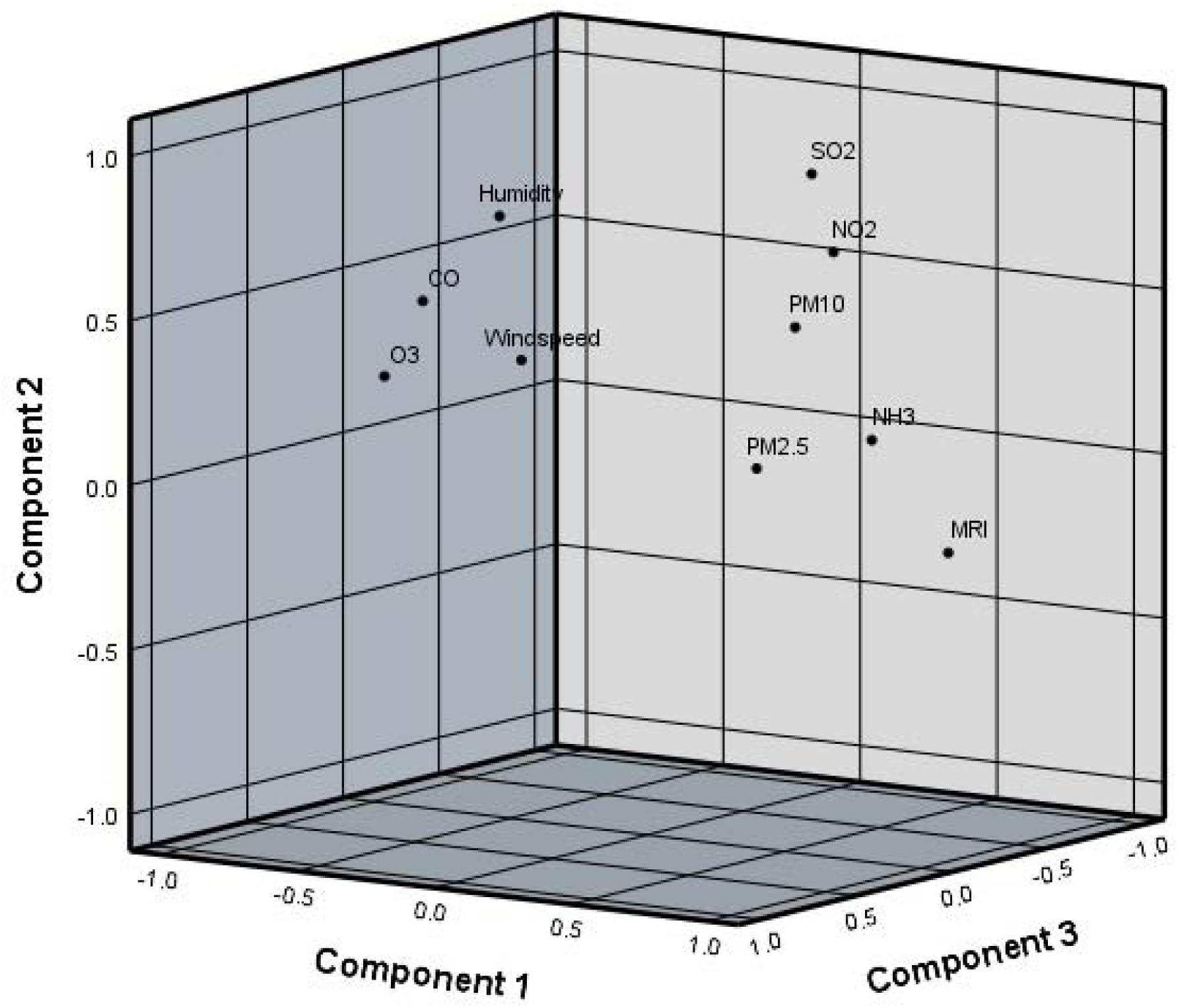
3-D view PCA of 10 parameters.

The cluster of PM_10_ & nitrogen dioxide contribute to health effects like, asthma in urban environments; lung inflammation and infections. PM_10_ & PM_2.5_ have adverse effect on human health effects like cough, asthma attacks & high blood pressure. The distance between the PM & SO_2_ depicted the decrease in childhood respiratory disease and all-age mortality caused by sulphur dioxide over the years.^23^ NH_3_ & MRI; the exposure to NH_3_ increases the level of intracellular polyamines leading to modifications in membrane permeability to different antibiotics as well as increased resistance to oxidative stress.^24^ O_3_, CO, RH & WS were inversely proportional to effect of other pollutants above, in urban environment during winter (Figure 5).^25^

**Figure 5.**
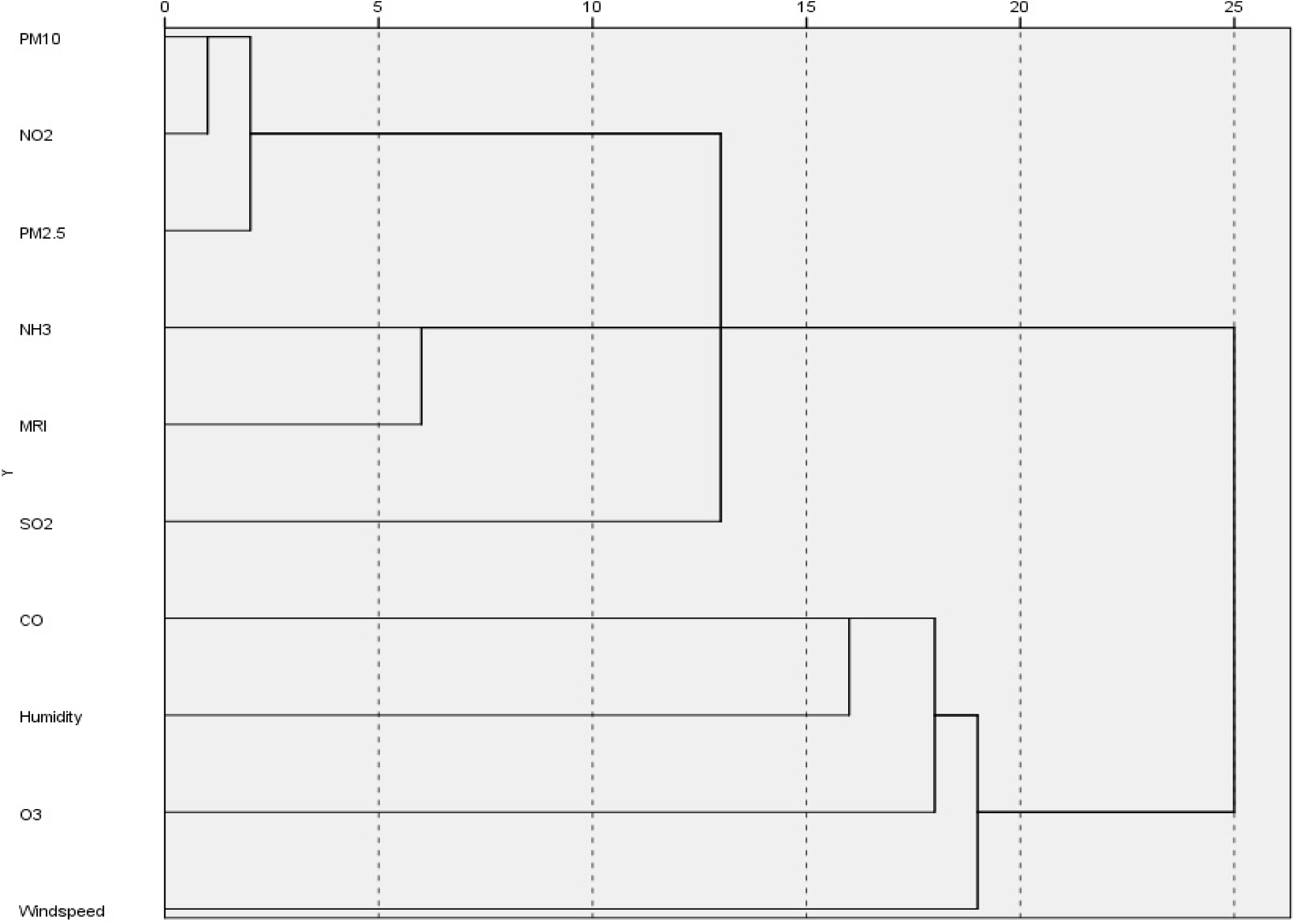
Dendrogram of 10 parameters.

AMP, NX & V relation depicted synergistic activity; AMP can replace with NX if the bacteria are beta-lactam resistant. E is bacteriostatic in nature; if the bacteria still multiply TR can be used to treat *S. pneumoniae* and *H. influenza.*^26,27^ COX *is* active against penicillinase-producing *Staphylococci spp*. & if the person is allergic to MET.^28^ CTR & C are bactericidal in nature and their synergistic activity is bacteriostatic. It’s active against *GBS* & *E. coli*.^29^ No TGC resistance was observed. TGC, lacking cross-resistance with other compounds, could represent an opportunity to reduce the intensity of selection for ESBL-producing organisms derived from the use of other antimicrobial agents (Figure 6).^30^

**Figure 6.**
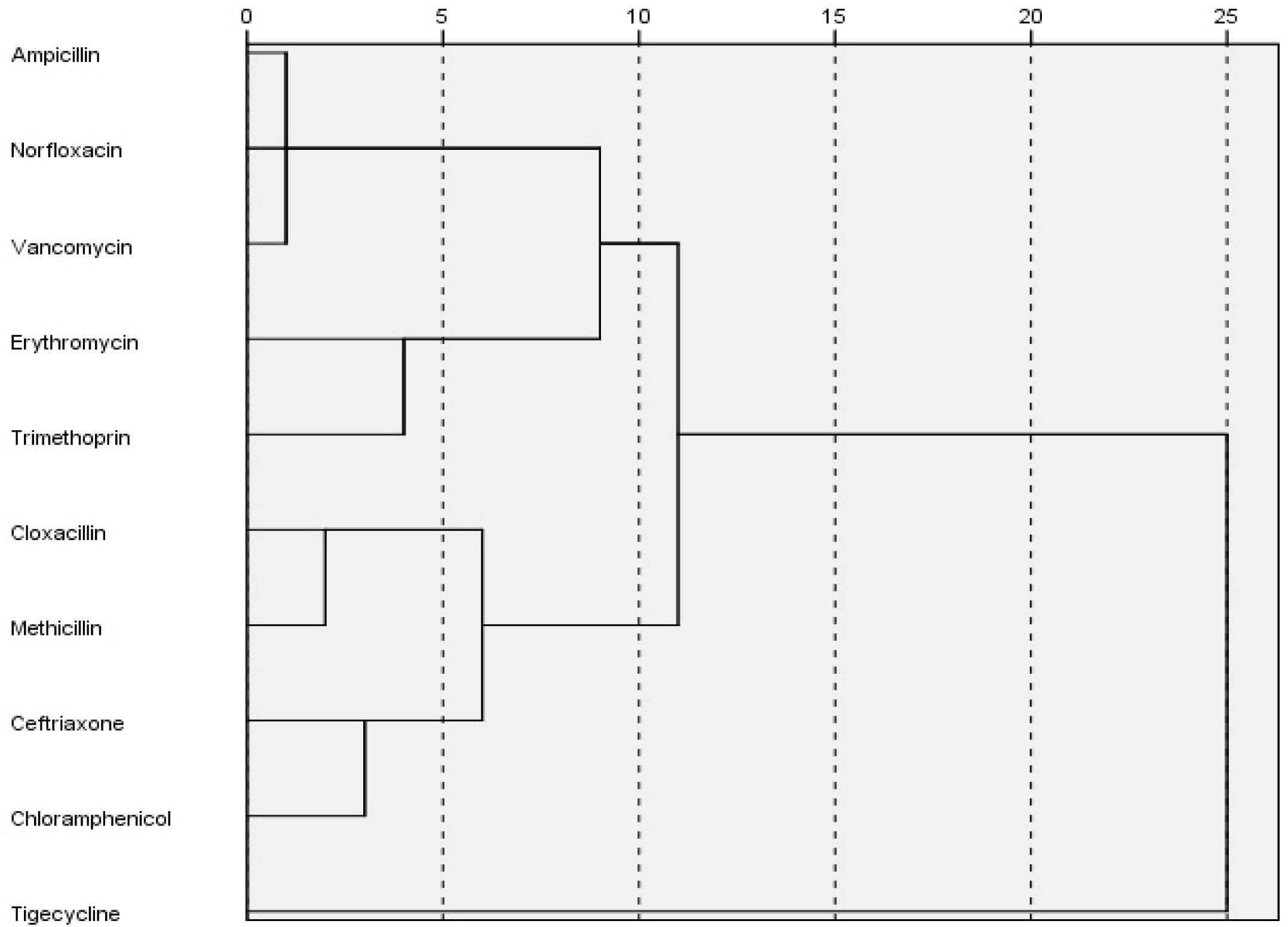
Dendrogram of 10 antibiotics.

The clustering of the locations is on the basis of physiochemical parameters and population. S5 & S6 locations are urban areas in Navi Mumbai (Table 1). The clustering of the locations was also dependent on the data (Table 2). The prominent pollutant of locations S1, S4 & S5 was PM_2.5_. The distance between S5 & S1 is lesser followed by S4. S2 & S7 have similar number of population (Table 1); they also had same levels of NH_3_ & RH (Table 2). S8 showed PM_10_ as the major pollutant and S2 had at similar levels. The locations which showed similar levels of PM_10,_ RH were S2 & S8; S3 was found to have distinct levels of RH (Figure 7).

**Figure 7.**
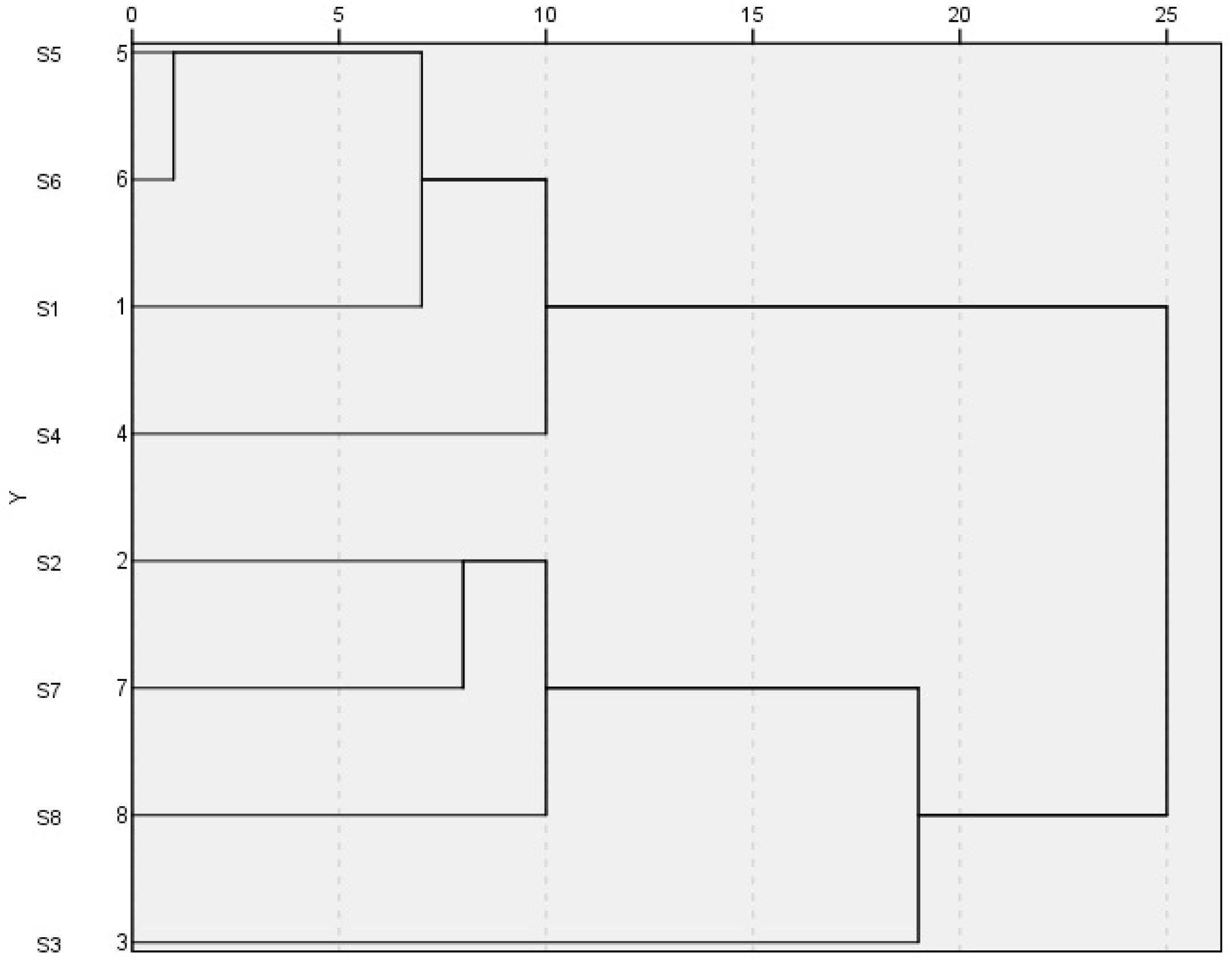
Dendrogram of 8 locations.

## Conclusion

The airborne microbiome enters into the atmosphere by aerosolization. The existence of microbes in the air creates ecological balance and also causes ill effects. The physiochemical interaction with these microbes helps to sustain them in harsh conditions and nutrient deficit environment. The bioaerosol are ubiquitous constituents of the atmosphere, as a large number of these particles are small sized microbes. Air pollution enhances the chances of health effects caused by particulate matter. Overall, the bacteria in outdoor air samples are predominantly derived from soil and plants sources. In this study, the concentration of bacteria was below average. The prevalence rate of the antibiotic resistant bacteria is higher. The results showed no correlation of the physiochemical parameters with multidrug resistant bacteria; may be because the airborne microbes originate from other sources. Therefore, it is essential to take preventive measures like sanitation and drinking clean water. This study showed that the airborne bacteria travel from different sources and come in contact with living organisms. It creates spread of pathogenic bacteria and may cause an infection.

COVID19 being a virus can be a better example of spread of infection through air droplets. The overuse of antibiotics, inappropriate disposal of antibiotics, presence of drugs in feces, etc. leads to antibiotic resistance.

In this pandemic period and lockdown, the air quality has improved. This shows that air pollutants have high impact not only on the air quality but also on the biodiversity.

## Data Availability

All data produced in the present work are contained in the manuscript

## Acknowledgement

The authors greatly acknowledge the director of SBB, DYP Prof. Debjani Dasgupta for allowing the lab space. Her continuous support and intellectual inputs were proved to be valuable for completion of the above study.

